# Increased levels of inflammatory molecules in blood of Long COVID patients point to thrombotic endotheliitis

**DOI:** 10.1101/2022.10.13.22281055

**Authors:** Simone Turner, Caitlin A Naidoo, Thomas J Usher, Arneaux Kruger, Chantelle Venter, Gert Jacobus Laubscher, M Asad Khan, Douglas B Kell, Etheresia Pretorius

**Author notes:** **Corresponding authors: Etheresia Pretorius**, Department of Physiological Sciences, Stellenbosch University, Private Bag X1 Matieland, 7602, SOUTH AFRICA, http://www.resiapretorius.net/, **Douglas B. Kell**, Department of Biochemistry and Systems Biology, Institute of Systems, Molecular and Integrative Biology, Faculty of Health and Life Sciences, University of Liverpool, L69 7ZB, UK.

## Abstract

The prevailing hypotheses for the persistent symptoms of Long COVID have been narrowed down to immune dysregulation and autoantibodies, widespread organ damage, viral persistence, and fibrinaloid microclots (entrapping numerous inflammatory molecules) together with platelet hyperactivation. Here we demonstrate significantly increased concentrations of Von Willebrand Factor, platelet factor 4,serum amyloid A, α-2antiplasmin E-selectin, and platelet endothelial cell adhesion molecule-1, in the soluble part of the blood. It was noteworthy that the mean level of α-2-antiplasmin exceeded the upper limit of the laboratory reference range in Long COVID patients, and the other 5 were significantly elevated in Long COVID patients as compared to the controls. This is alarming if we take into consideration that a significant amount of the total burden of these inflammatory molecules has previously been shown to be entrapped inside fibrinolysis-resistant microclots (thus decreasing the apparent level of the soluble molecules). We also determined that by individually adding E-selectin and PECAM-1 to healthy blood, these molecules may indeed be involved in protein-protein interactions with plasma proteins (contributing to microclot formation) and platelet hyperactivation. This investigation was performed as a laboratory model investigation and the final exposure concentration of these molecules was chosen to mimic concentrations found in Long COVID. We conclude that presence of microclotting, together with relatively high levels of six inflammatory molecules known to be key drivers of endothelial and clotting pathology, points to thrombotic endotheliitis as a key pathological process in Long COVID. This has implications for the choice of appropriate therapeutic options in Long COVID.

**SENTENCE SUMMARY:** The presence of fibrinaloid microclots and multiple inflammatory molecules in the soluble part of blood points to thrombotic endotheliitis as a key pathological process in Long COVID.

## INTRODUCTION

Post-Acute Sequelae of COVID-19 (PASC) or Long COVID is a major global health burden with its symptoms significantly impacting physical and cognitive function, health-related quality of life, and participation in society (*1-9*). The mechanisms for the persistent symptoms can be narrowed down to 1) viral persistence, 2) microclotting and platelet hyperactivation, 3) autoantibodies, 4) immune dysregulation, 5) widespread organ damage, and the 6) reactivation of dormant viruses (*9-12*).

SARS-CoV-2 (COVID-19) viral remnants including RNA and spike protein have been identified in patients with Long COVID. Increases in antibody responses directed against non-SARS-CoV-2 viral pathogens, particularly Epstein-Barr virus, have been noted and are likely to be related to reactivation of dormant viruses (*13, 14*). There is also increasing evidence that autoantibodies may be involved in the lingering symptoms of individuals with Long COVID (*15, 16*). We have demonstrated that individuals with Long COVID have a significant fibrin amyloid microclot load in their circulation (*2, 10, 17*). These clots can be induced in normal plasma by (recombinant) SARS-CoV-2 spike protein (*18*), and are resistant to fibrinolysis (*10, 19*). Microclots are microscopic (fibrin-amyloid-containing) clots present in the plasma of recruited individuals (*10, 19*). Entrapped in these fibrinolysis-resistant microclots are numerous inflammatory molecules, including alpha 2-antiplasmin (α2AP), various fibrinogen chains, Von Willebrand factor (VWF), platelet factor 4 (PF4), Serum Amyloid A (SAA), as well as numerous antibodies (*10, 19*). If the coagulopathy present in the acute phase of the disease, is not adequately treated, it is likely that both tissue hypoxia and impaired oxygen exchange may linger for months (*10, 11, 17, 19, 20*). Recently it was noted that the incidence of vascular events remains elevated up to 49 weeks after COVID-19 diagnosis (*21*). The published evidence so far points to widespread endothelial inflammation (*22-24*), and the underlying causes are a failed (i.e. inadequate) fibrinolytic system and persistent coagulopathy (*9*).

Although characterizing the inflammatory molecule content of the fibrinolytic-resistant microclots is of considerable interest and value (*2, 10, 17, 20*), patients desperately need more readily accessible diagnostic markers at general pathology laboratories. In this paper we focus on identifying biomarkers in the soluble fraction of blood that could be used as a suite to confirm the widespread endothelial damage and coagulation pathologies see in Long Covid patient. The soluble molecules that were chosen for this study, were chosen from previously published research that found these molecules entrapped inside solubilized microclots, as well as two additional molecules. The previously identified molecules include VWF, PF4, SAA and α-2AP, with the two new markers, that are well known endothelial damage markers, include endothelial-leukocyte adhesion molecule 1 (E-selectin), and the platelet endothelial cell adhesion molecule1 (PECAM-1).

### Von Willebrand Factor

Following damage to the endothelial layers, VWF is released by endothelial cells (*25*) and megakaryocytes; it mediates platelet adhesion to the damaged vascular surface, and the aggregation of platelets (*26, 27*). We included VWF as it is stored in endothelial Weibel-Palade bodies (*28*) and platelet α-granules (*29*), and is also secreted during platelet activation and endothelial damage (*30, 31*). Back in 2020, we and other researchers discussed the importance of VWF as a disease marker in acute COVID-19 (*22, 32-36*). It has also been noted that VWF/ADAMTS-13 imbalance persists in endothelial cell activation and angiogenic disturbance in Long COVID (*37*).

### Platelet Factor 4

Another important inflammatory molecule driving coagulation and platelet pathology, is PF4 (*38-40*). Platelets produce and store PF4 in platelet α-granules bound to the glycosaminoglycan (GAG) chains of serglycin (*41*). Heparin can bind to PF4 and thereby promotes PF4 aggregation, resulting in the formation of PF4/heparin complexes, which have antigenic properties (*38*). PF4 and VWF may also form complexes that are involved in thrombus propagation (*39*).

### α-2-antiplasmin

Central to the persistent nature of microclots in Long COVID (in addition to the simple resistance to proteolysis of amyloid-like proteins (*42*) entrapping inflammatory molecules), is a plasmin and antiplasmin imbalance in individuals with Long COVID. We have found that α-2AP is trapped inside microclots (*10, 19*). This molecule prevents the fibrinolytic system from functioning optimally and impedes the digestion of pathological thrombi that cause microvascular thrombosis, thereby increasing the risk of thromboembolic events (*10, 19*). High blood levels of α-2AP, an ultrafast, covalent inhibitor of plasmin-increase the risk of a poor outcome in cardiovascular diseases (*43*).

### Serum amyloid A

We also included, as part of our suite of soluble inflammatory markers, the more general inflammatory marker, SAA. The human SAA protein family comprises the acute phase SAA1 and SAA2, involved in innate and adaptive immunity and the constitutive SAA4 (*44, 45*). SAA activates the complement system and the nucleotide-binding domain, leucine-rich repeat (NLR) family pyrin domain containing 3 (NLRP3) inflammasome, alters vascular function, affects HDL function, and increases thrombosis (*46*).

### Endothelial factors

As endothelial dysfunction is central in persistent symptoms of Long COVID, we also chose to include two well-known endothelial damage markers, namely, CD62 antigen-like family member E (CD62E) a commonly known as E-selectin (sometimes also called endothelial-leukocyte adhesion molecule 1 (ELAM-1), and the platelet endothelial cell adhesion molecule-1 (PECAM-1). E-selectin is a glycoprotein cell adhesion receptor that is expressed on endothelial cells activated by cytokines (*47*), and can be used as a marker to evaluate endothelial dysfunction (*48, 49*). Platelet endothelial cell adhesion molecule-1 (PECAM-1), also referred to as CD31, functions as an inhibitory receptor in circulating platelets and is highly expressed at endothelial cell-cell junctions, where it functions as an adhesive stress-response protein to both maintain endothelial cell junctional integrity (*50*). As PECAM-1 is also expressed on platelets and leukocytes, it is at the nexus of thrombosis and inflammation (*51*).

In addition to determining if E-selectin and PECAM-1 are upregulated in the soluble part of the blood, we also wished to determine if these they may indeed be involved in protein-protein interactions with plasma proteins to cause the formation of (amyloid) protein misfolding (resulting in microclot formation). Furthermore, we investigated if these two molecules can interact with and bind to platelet receptors to cause platelet hyperactivation. This part of the investigation was planned as a laboratory model investigation and the final exposure concentration was chosen to mimic concentrations found in Long COVID.

## MATERIALS AND METHODS

### Ethical clearance

Ethical clearance for the study was obtained from the Health Research Ethics Committee (HREC) of Stellenbosch University (South Africa) (references: B21/03/001_COVID-19, project ID: 21911 (long COVID registry) and N19/03/043, project ID 9521 with yearly re-approval). Participants were either recruited via the Long COVID registry or identified from our clinical collaborator’s practice. The experimental objectives, risks, and details were explained to volunteers and informed consent was obtained prior to blood collection. Strict compliance to ethical guidelines and principles of the Declaration of Helsinki, South African Guidelines for Good Clinical Practice, and Medical Research Council Ethical Guidelines for Research were kept for the duration of the study and for all research protocols.

### Patient demographics

#### Participant identification for ELISA analysis

For the enzyme-linked immunosorbent assay (ELISA) analysis, we randomly selected a subset of stored Long COVID and healthy samples, where we previously confirmed entrapped inflammatory molecules using proteomics analysis (*10*) (see Table 1). The sample included blood collected from 15 healthy individuals to serve as controls (1 male; 14 females; median age 42 [27-61]) and 25 Long COVID patients (9 males; 16 females; median age 52 [22-76]). Healthy participants did not smoke, did not suffer from coagulopathies, and were not pregnant. The questionnaire contained information regarding their age, gender, pre-existing comorbid conditions, chronic medication, information regarding the onset, diagnosis, and severity of their acute COVID-19 illness, self-reported long COVID symptoms, as well as their COVID-19 vaccination history.

**Table 1:** Sample demographics and considerations.

| Previously stored PPP and serum |  |
| --- | --- |
| Mean age [and standard deviation (SD)] of controls and Long COVID |  |
| Controls (n=15) | 42 (± 8) |
| Long COVID (n=25) | 52 (±14) |
| Vaccination status of controls and Long COVID patients before blood collection |  |
| Percentage of controls vaccinated (n=15) | 33% |
| Percentage of Long COVID/PASC patients vaccinated (n=25) | 24% |
| Co-morbidities of Long COVID patients (n=25) |  |
| Co-morbidity | % In 25 Long COVID patients |
| Hypertension | 36% |
| Hyperlipidaemia | 40% |
| Type 2 Diabetes Mellitus | 12% |
| Rosacea | 0% |
| Thrombosis (previous blood clots) | 8% |
| Cardiovascular disease | 8% |
| Psoriasis | 4% |
| Rheumatoid arthritis | 4% |
| Previous stroke | 0% |
| Previous myocardial infarction | 4% |
| COPD | 4% |
| Cancer | 12% |
| Gout | 4% |
| **Laboratory study of healthy whole blood and platelet poor plasma** |  |
| **Mean age [and standard deviation (SD)] of controls** |  |
| Mean age [and standard deviation (SD)] (n=6) | 41( ±15) |
| **Vaccination status of controls before blood collection** |  |
| Percentage of controls vaccinated (n=6) | 100% |

#### Participant identification for blood laboratory model

Blood was collected from six healthy individuals [1 male and 5 females; 41(±15)]. We used these blood samples to study the effects of two inflammatory molecules that originate from damaged endothelial cells. These individuals were all previously vaccinated. Two of the controls developed pericarditis after their vaccination but was effectively treated and had no clinical features of vaccine injury at time the blood was drawn (more than a year later).

### Blood sample collection and sample preparation

Either a qualified phlebotomist or medical practitioner drew blood samples. To obtain platelet poor plasma (PPP), whole blood (WB) from sodium citrate tubes was centrifuged at 3000xg for 15 min at room temperature and the supernatant PPP samples were collected in 1.5 mL Eppendorf tubes and stored at –80 °C. To obtain plasma, WB from rapid serum tubes was centrifuged at 3000xg for 15 min at room temperature and the supernatant plasma samples were collected in 1.5 mL Eppendorf tubes and stored at –80 °C.

### Enzyme-linked immunosorbent assays (ELISA)

Enzyme-linked immunosorbent assay (ELISA) were performed on all the biomarkers except VWF. The ELISA recognizes and quantitatively measures the molecule of interest in samples using the Sandwich-ELISA principle. The 96 well plate that is provided in the kit is pre-coated with an antibody specific to the molecule of interest. The samples were then diluted according to each ELISA kit detection range. After the samples were correctly diluted, 100μL of the diluted samples and standards were added to the wells and combined with the specific antibody. Next, the biotinylated detection antibody specific for the molecule of interest and Avidin-Horseradish Peroxidase (HRP) conjugate were added successively to each micro plate well and incubated. After the wash step, the substrate solution is added to each well. The enzyme-substrate reaction is terminated by the addition of stop solution. The optical density was measured spectrophotometrically at a wavelength of 450 ± 2 nm with the BioTek Synergy HTX Multi-Mode Microplate Reader (Santa Clara, United States). The concentrations were calculated using Graphpad Prism 8 (version 9.1.1).

Serum Amyloid A was measured using Abcam’s Human SAA ELISA Kit (ab100635) using a dilution factor of 500x. α-2 antiplasmin (α-2AP) was measured using Abcam’s Human alpha-2 antiplasmin SimpleStep ELISA^®^ Kit (ab254502) using a dilution factor of 20 000x. Platelet factor 4 was measured using Elabscience’s Human PF4 ELISA Kit (E-EL-H6107) using a dilution factor of 10x. E-selectin was measured using Elabscience’s E-selectin ELISA Kit (E-EL-H0876) using a dilution factor of 20x. Platelet endothelial cell adhesion molecule (PECAM) was measured using Elabscience’s PECAM ELISA Kit (E-EL-H1640) using a dilution factor of 50x.Only PECAM-1 and E-selectin were measured using serum, the rest of the molecules were measured used PPP.

### Von Willebrand Factor antigen

VWF was analysed by a pathology service laboratory known as PathCare, located in Mediclinic Stellenbosch, South Africa. We chose to test for VWF % antigen at Pathcare as VWF is the only molecule out of the six inflammatory molecules that is a standard test that can be performed in a pathology laboratory. This also makes the test more assessable to patients and doctors.

For sample analysis, Siemens VWF Ag reagent was used for the immunoturbidimetric determination, using the Sysmex CS2500 instrument. Approximately 0.5mL PPP was utilized for the analysis. The Sysmex CS2500 instrument was calibrated according to standard protocol and the facility is accredited in accordance with the international ISO 15189:2012 standard.

### Fluorescence microscopy

#### Platelet poor plasma (PPP) analysis of the Long COVID samples

Fluorescence microscopy to study the presence of microclot formation in PPP was performed on some of our samples as described in our previous papers (*52*). Stored PPP samples were exposed to Thioflavin T (ThT), a fluorogenic dye that binds to amyloid protein (*42*) after reached room temperature. A final concentration of 0.005mM was used (Sigma-Aldrich, St. Louis, MO, USA). Plasma was exposed for 30 minutes (protected from light) at room temperature (*42, 53-56*), wgereafter 3µL stained PPP was placed on a glass slide and covered with a coverslip.

#### Healthy blood samples with added E-selectin and PECAM-1

Whole blood drawn from six healthy participants was centrifuged and PPP and haematocrit samples were prepared as described in the above paragraphs. Microclot formation and platelet activation were studied in the naïve samples. For this analysis, the PPP was exposed to ThT as described in the previous paragraph. The remaining haematocrit samples were retained and were incubated for 30 minutes at room temperature, with the two fluorescent markers, CD62P (PE-conjugated) (platelet surface P-selectin) (IM1759U, Beckman Coulter, Brea, CA, USA) and PAC-1 (FITC-conjugated) (340507, BD Biosciences, San Jose, CA, USA) (17). CD62P is a marker for P-selectin that is either found on the membrane of platelets or inside them. PAC-1 identifies platelets through marking the glycoprotein IIβ/IIIα (gpIIβ/IIIα) on the platelet membrane.

Both the PPP and the haematocrit samples from the 6 healthy individuals were then also exposed for 30 minutes, at room temperature, to E-selectin and PECAM-1 at a final concentration of 30 ng.ml^-1^. Recombinant Human E-Selectin/SELE Protein (His Tag) (Catalog No. PKSH032403) and Recombinant Platelet/Endothelial Cell Adhesion Molecule (PECAM-1) (Catalog No. RPA363Hu01) was purchased from Biocom Africa (Centurion, South Africa). After the 30 minute exposure time, the E-selectin and PECAM-1-exposed, PPP samples were incubated with ThT. The E-selectin and PECAM-1-exposed haematocrit samples were also incubated with the two platelet markers, CD62P and PAC-1 (as described in the above paragraphs).

To view ThT exposed PPP samples, the excitation wavelength was set at 450nm to 488nm and the emission wavelength at 499nm to 529nm. The excitation wavelength for the hematocrit samples exposed to PAC-1 was set at 450nm to 488nm and the emission at 499 to 529nm and for the CD62P marker the excitation wavelength was set at 540nm to 570nm and the emission 577nm to 607nm. All samples were viewed using a Zeiss Axio Observer 7 fluorescent microscope with a Plan-Apochromat 63x/1.4 Oil DIC M27 objective (Carl Zeiss Microscopy, Munich, Germany).

### Statistical analysis

GraphPad Prism software (version 9.1.1) was used to perform statistical analysis on all data. To determine whether data is normally distributed, the Shapiro-Wilk normality test was applied. Parametric data was analysed using an unpaired, one-tailed t-tests with the Welch’s correction in order to test for statistical significance. For non-parametric data, an unpaired, one-tailed Mann-Whitney test was utilized to test for statistical significance. Statistical significance was established at p< 0.05 (* = p<0.05; ** = p<0.01). Data is either represented as either mean ± standard deviation (parametric data) or as median ± [Q1-Q3] (non-parametric data).

## RESULTS

### Enzyme-linked immunosorbent assays (ELISAs)

Results of our five ELISA experiments (SAA, α-2AP, PF4, E-selectin and PECAM-1) and VWF measured at a pathology laboratory, are shown in Figure 1. All six inflammatory molecules were significantly upregulated in the soluble part of the blood that was measured (See Table 2).

**Table 2:** Inflammatory molecule concentration [or % antigen in the case of Von Willebrand Factor (VWF)] data of 15 controls and 25 Long COVID patients. Parametric data is expressed as Mean (SD), and non-parametric data as Median(Q1-Q3). **Abbreviations:** SD: standard deviation.

| Inflammatory molecule | Reference range | Controls (n=15):<br>Mean (SD) OR<br>Median (Q1-Q3) | Long COVID(n=25):<br>Mean (SD) OR<br>Median (Q1-Q3) | Unit | P-value |
| --- | --- | --- | --- | --- | --- |
| **SAA** | 0-10 | 5.3(1.9-8.0) | 6.9(4.8-17.25) | mg.L<sup>-1</sup> | **\*p<0.05** |
| **α-2AP** | 60-80 | 71.73(±18.48) | 90.28(±11.31) | mg.L<sup>-1</sup> | **\*\*p<0.01** |
| **PF4** | 197-1390 | 484.2(412.8-526.8) | 572.4(430.6-779.9) | ng.ml<sup>-1</sup> | **\*p<0.05** |
| **VWF** | 55.9 - 161.6 | 76.6(±28.03) | 104.8(±60.1) | % | **\*p<0.05** |
| **E-selectin** | 8.5-26 | 10.26(±3.07) | 13.86(±5.4) | ng.ml<sup>-1</sup> | **\*\*p<0.01** |
| **PECAM-1** | 5.3-15 | 8.19(7.06-10.08) | 10.27(8.32-11.45) | ng.ml<sup>-1</sup> | **\*p<0.05** |

**Figure 1:**
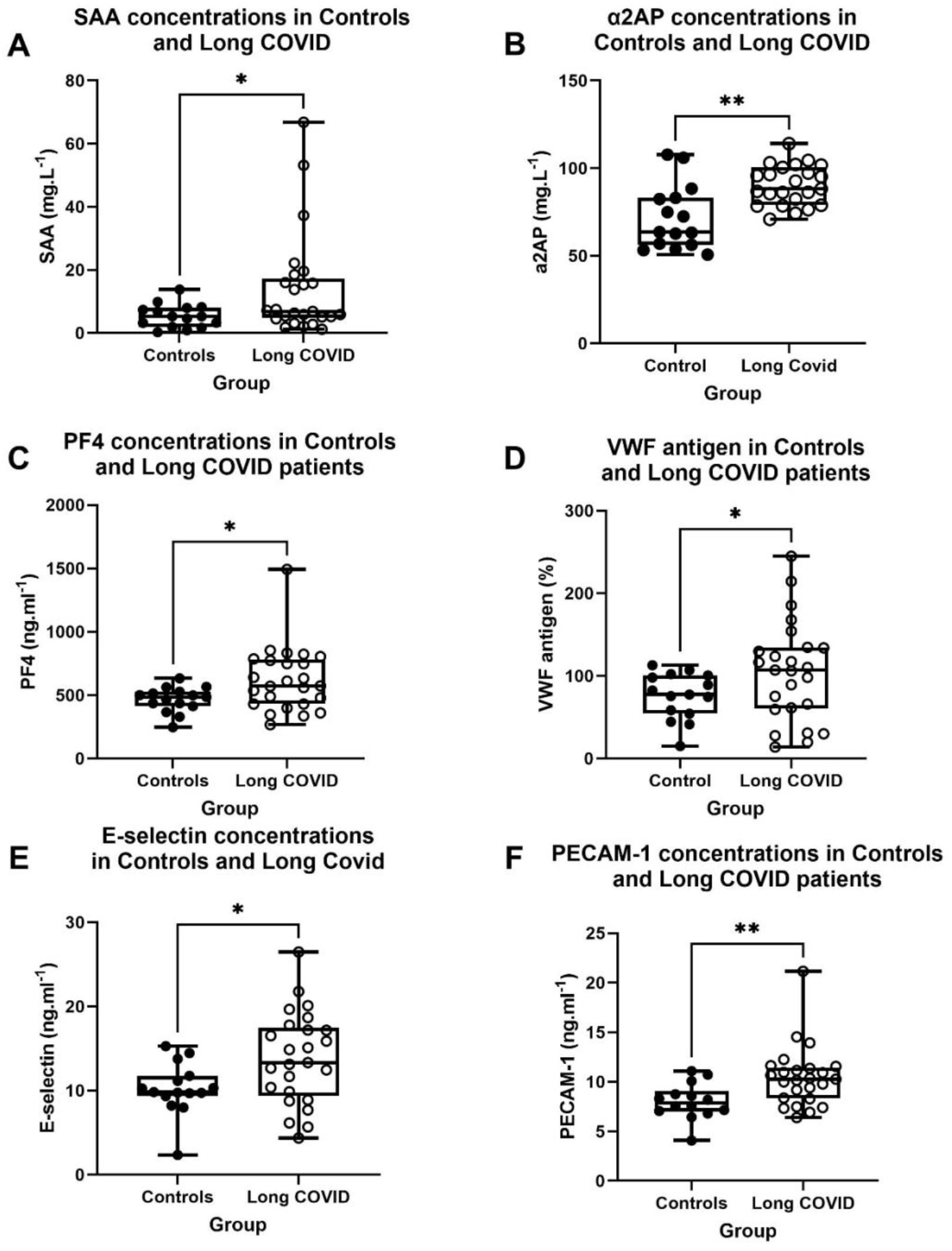
Inflammatory molecule concentration [or % antigen in the case of Von Willebrand Factor (VWF)] in controls and Long COVID within systemic circulation. **(A)** SAA-, **(*p<0.05), (B)** α2AP-**(**p<0.01), (C)** PF4-**(*p<0.05)** and **(D)** VWF concentrations in controls and Long COVID using PPP **(*p<0.05). (E)** E-selectin-and **(**p<0.01). (F)** PECAM-1 concentrations in controls and Long COVID using serum **(*p<0.05). Abbreviations:** SAA: Serum Amyloid A, α-2AP: α-2 antiplasmin, PF4: Platelet factor 4, VWF: Von Willebrand Factor, E-selectin: endothelial-leukocyte adhesion molecule 1, PECAM-1: Platelet endothelial cell adhesion molecule-1, PPP: Platelet poor plasma.

### Fluorescence microscopy

Figure 2 show representative micrographs of the Long COVID samples. These results are in line with previous reports, with major microclot formation noted in these individuals. Figures 3 and 4 show representative micrographs of healthy PPP and haematocrit samples exposed to either E-selectin or PECAM-1, that resulted in considerable plasma protein aggregation and platelet activation after addition of both these two inflammatory molecules.

**Figure 2:**
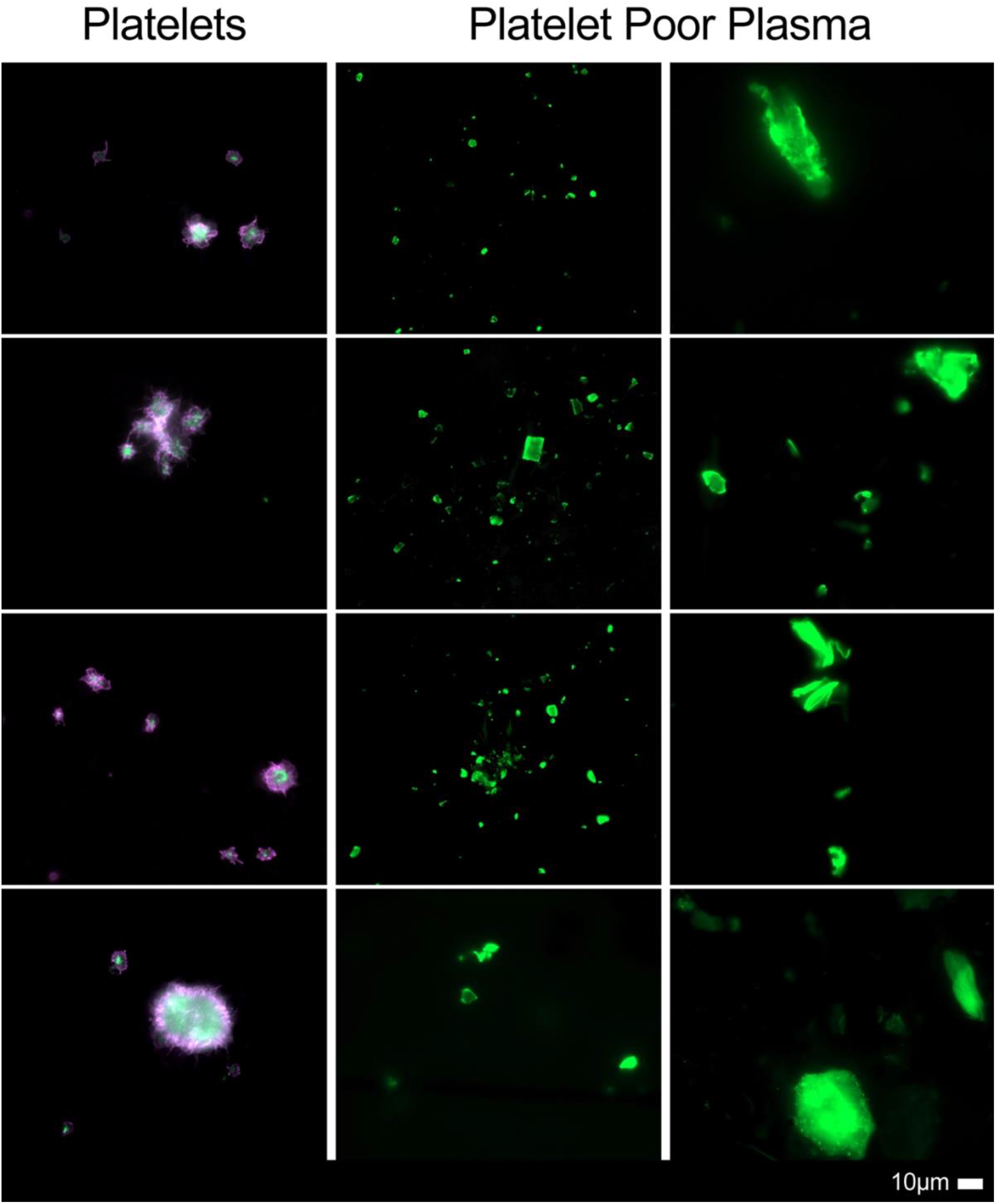
Micrographs of platelets and platelet poor plasma (PPP) [with added thioflavin T (ThT)] from patients with Long COVID/PASC, showing widespread microclot formation.

**Figure 3:**
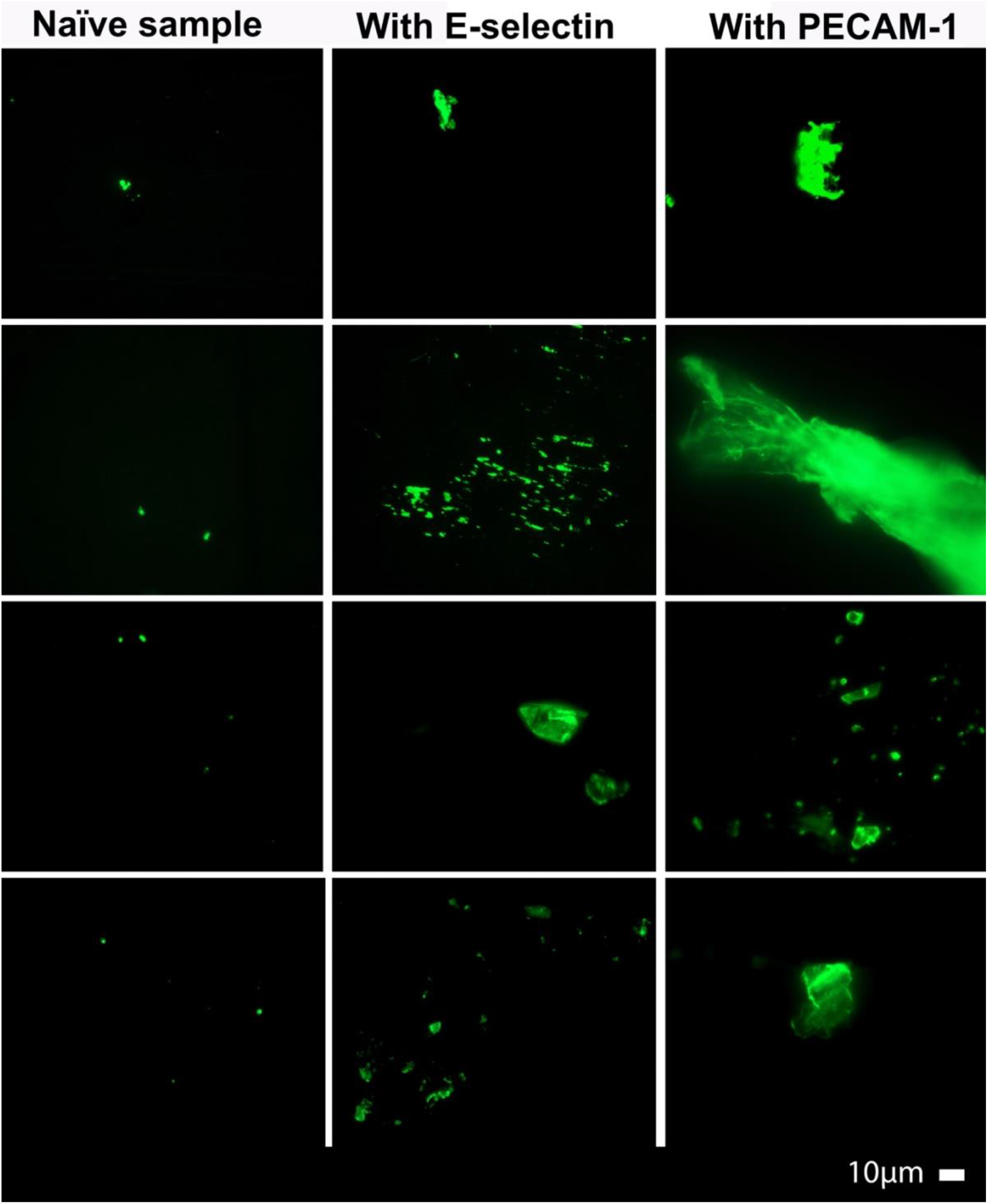
Micrographs of platelet poor plasma (PPP) from healthy individuals exposed to either E-selectin (final exposure concentration of 30 ng.ml^-1^) or PECAM-1 (final exposure concentration 30 ng.ml^-1^) followed by exposure to thioflavin T (ThT), showing the effects of the two molecules on the plasma.

**Figure 4:**
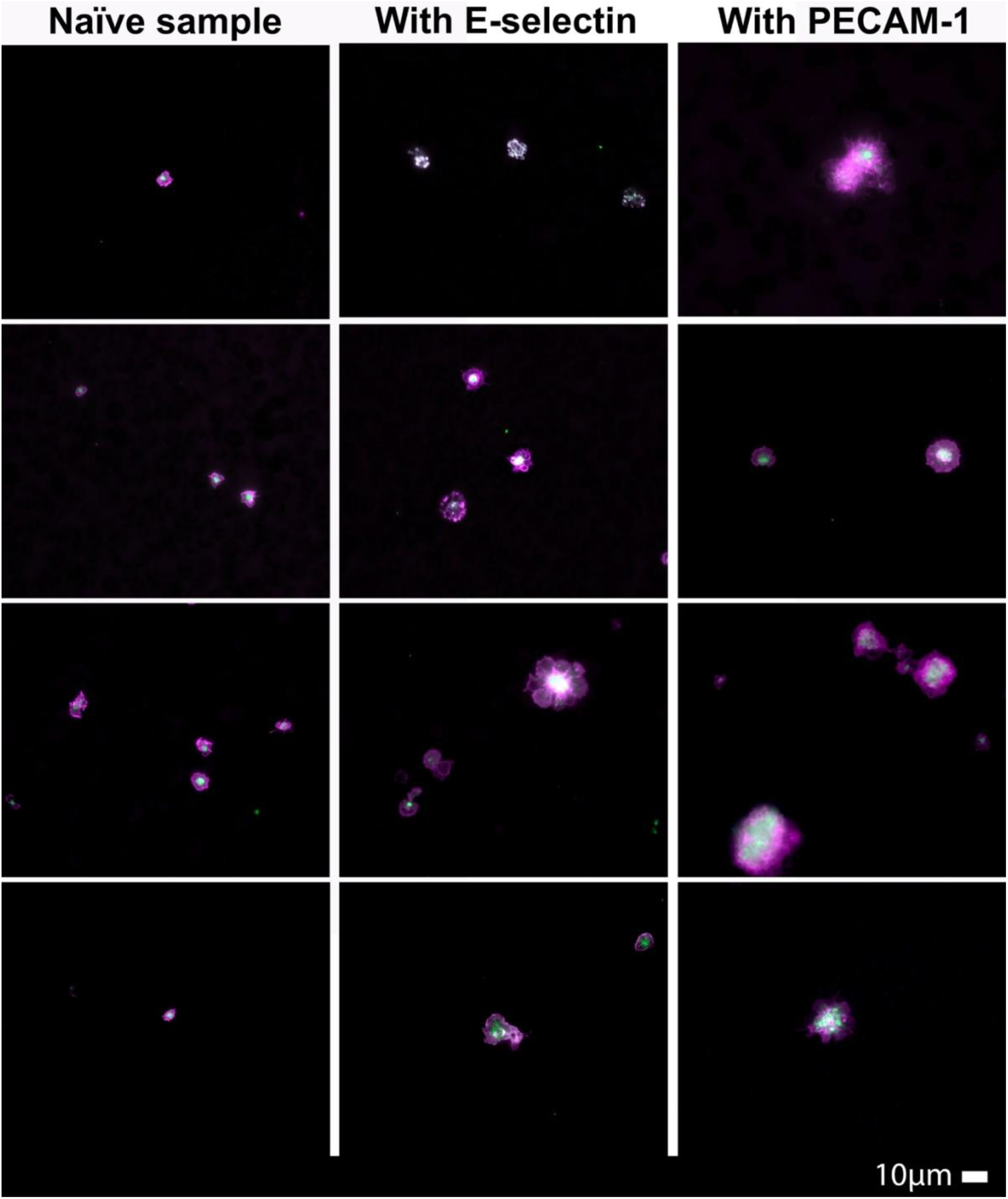
Micrographs of haematocrit from healthy individuals exposed to either E-selectin (final exposure concentration of 30 ng.ml^-1^) or PECAM-1 (final exposure concentration of 30 ng.ml^-1^) followed by exposure two platelet markers, PAC-1 (green) and CD62P (pink).

## DISCUSSION

We have previously determined that various inflammatory molecules are entrapped inside fibrinolysis-resistant microclots in individuals with Long COVID (*10, 19*). We undertook proteomics analysis and with a double digestion method to liberate these molecules. We identified, amongst others, VWF, α-2AP, SAA and PF4 entrapped inside these microclots (*10, 19*). Interestingly, Captur and co-authors more recently also noted that the plasma proteomic signature predicts who will get persistent symptoms following SARS-CoV-2 infection (*57*). They also found that SAA and α-2AP were upregulated in their cohorts (*57*). These molecules and similar such molecules are also well-known to activate platelets, by binding to their receptors (*58, 59*). Both microclots and hyperactivated platelets, may interact with damaged endothelial cells and further perpetuate widespread vascular damage. In addition, E-selectin and PECAM-1 are well-known markers of endothelial damage, that was also analysed in this paper.

In the current paper, we have chosen to determine if the concentrations of the above 6 inflammatory markers are also upregulated in the soluble part of blood (serum and plasma). We have also shown (Figures 3 and 4) that both E-selectin and PECAM-1 can trigger considerable microclot formation as well as platelet hyperactivation similar to that previously noted in samples from individuals suffering from Long COVID (See Figure 2).

To understand how these six molecules may be central in perpetuating widespread endothelial damage and persistent coagulation pathology noted in Long COVID, we need to turn our attention to the physiological pathways that are affected by them. We focus on receptor activation pathways in endothelial cells and platelets.

### Von Willebrand Factor

VWF is well-known inflammatory molecule that is widely used as a marker of inflammation and vascular damage (*25*), and is commonly available in pathology laboratories. We have found VWF to be trapped inside microclots and also upregulated in the soluble part of the plasma (*10, 19*). When in circulation, it binds to two main receptors on the platelet membrane, including, GPIbα (in the GPIb-IX-V complex) and integrin αIIbβ3 (in the GPIIb-IIIa complex) (*60*) (see Figure 5). When VWF binds to GPIbα in the GPIb-IX-V complex, tyrosine protein kinases LYN and FYN are activated (*61*). When these kinases are activated, they promote tyrosine phosphorylation of the immunoreceptor tyrosine-based activation motif (ITAM) on the FcRγ or FcγRIIα (*61*). Once the GPIb-IX-V complex is engaged, it activates intracellular signalling events that instigate platelet activation and aggregation through the αIIbβ3 integrin (*61*). This also allows for more VWF to bind to αIIbβ3 as binding of VWF to GPIb-IX-V further upregulates αIIbβ3 integrin affinity (*62*). When additional VWF molecules bind to αIIbβ3, platelet adhesion and platelet aggregation are upregulated. This ultimately contributes to thrombus formation by binding to fibrinogen, which is mediated via different pathways (*62*) (see Figure 5). Whilst the fundamental role of VWF is to activate platelets, VWF can also bind to endothelial cells via the integrin αvβ3 receptor (*63*). An important function of VWF is to ensure endothelial cell adhesion, migration, proliferation, differentiation, and survival (Bryckaert et al., 2015). In addition, when VWF binds to the αvβ3 integrin on endothelial cells, it encourages Weibel-Palade bodies to secrete additional VWF into the circulation (*64*). The αvβ3 receptor on endothelial cells also binds fibrinogen and fibronectin (*63*).

**Figure 5:**
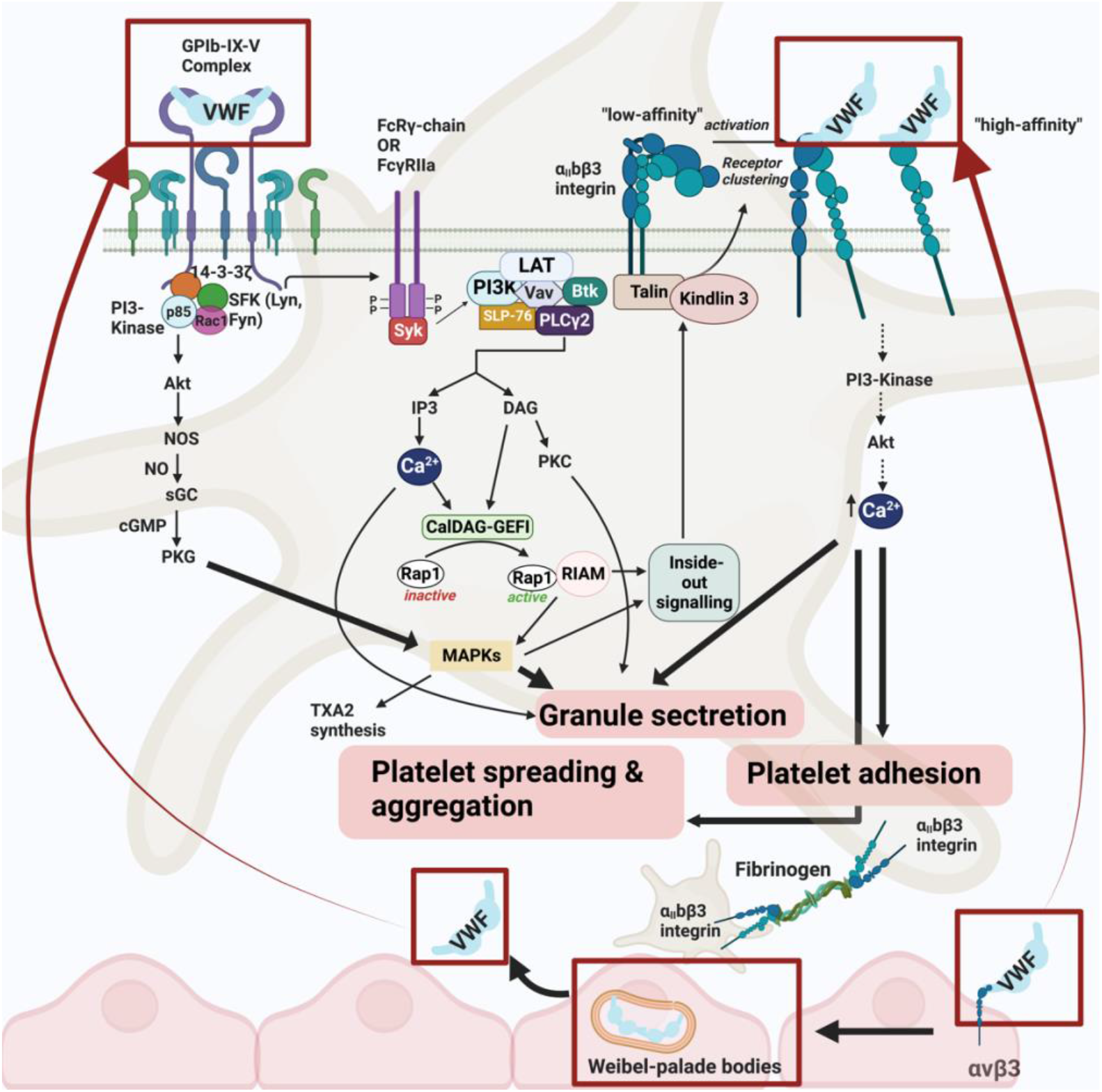
The receptors and signalling pathways of Von Willebrand factor (VWF) in platelets and endothelial cells. VWF binds to the GPIb-IX-V complex leading to integrin αIIbβ3 activation. Subsequently, various pathways are activated that ultimately lead to granule secretion, platelet adhesion, platelet spreading, and platelet aggregation that promote clot formation. VWF may also bind to endothelial cells via the αvβ3 integrin, which causes Weibel-palade bodies within endothelial cells to release VWF into circulation. **Abbreviations:** ADP, adenosine triphosphate; Btk, Bruton tyrosine kinase; CalDAG-GEFI, Ca^2+^-dependent guanine nucleotide exchange factor; DAG, diacylglycerol; IP3, inositol triphosphate; LAT, linker of activated T-cells; MAPK, mitogen-activated protein kinase; NO, nitric oxide; NOS, nitric oxide synthase; PI3K, phosphoinositide 3; PLC-γ2, phospholipase C-γ2, PKC, protein kinase C; PKG, protein kinase G; Rac 1, Ras-related C3 botulinum toxin substrate 1; Rap1, Ras-related protein 1; RIAM, Rap1-GTP-interacting adaptor molecule; SFK, Src family kinases; SLP-76, SH2 domain-containing leukocyte phosphorylation of 76 kDa; SYK, spleen tyrosine kinase; sGC, soluble guanine cyclase; TXA2, thromboxane A2; VWF, Von Willebrand factor. This image was created with BioRender (https://biorender.com/).

### Platelet Factor 4

We also noted a significant upregulation of PF4 in our Long COVID samples. PF4 is released from the α-granules of activated platelets as a complex with a chondroitin sulfate proteoglyan carrier (*65, 66*). After secretion from platelets, PF4 quickly transfers to higher affinity heparan sulfate on endothelial cells where it inhibits local antithrombin activity and promotes coagulation (*41*). When heparin-induced thrombocytopenia antibodies bind to PF4/heparin complexes, these immune complexes can activate platelets via the FcγIIα receptor, further promoting thrombin generation and increased coagulation (*67*). Interestingly, PF4 does not directly bind to endothelial cells, but rather enhances platelet–neutrophil interactions that activate the vascular endothelium (Walenga et al., 2004) (see Figure 6).

**Figure 6:**
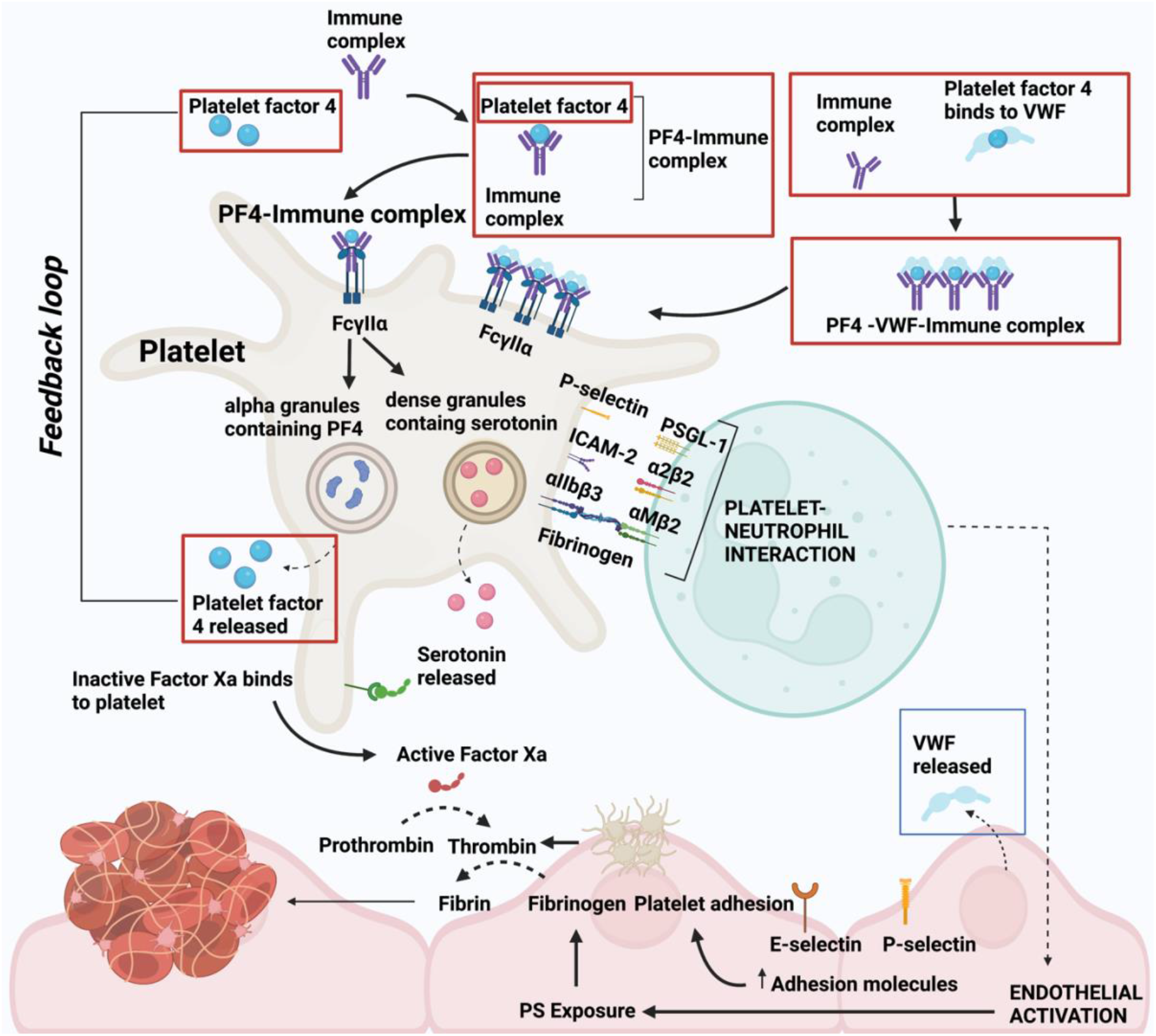
Platelet factor 4 and its receptor and signalling pathways in platelets and endothelial cells. PF4 binds to an immune complex and thereby binds to the FcγIIα receptor on platelets. PF4 can also bind to VWF and subsequently form larger VWF strands. This PF4-VWF-Immune complex can also bind to the FcγIIα receptor on platelets. Once bound to platelets, it causes alpha granules to release PF4, and dense granules to release serotonin. This causes further platelet activation and aggregation. Subsequently, neutrophils are recruited and bind to platelets via various receptors. The platelet-neutrophil interactions cause endothelial activation and an increase in expression of adhesion molecules and inflammatory molecules such as VWF. Endothelial activation also promotes PS Exposure and platelet adhesion to the endothelium, which promotes the production of thrombin. Thrombin then converts fibrinogen into fibrin, increasing the risk of clotting pathology. **Abbreviations:** PF4: platelet factor 4, VWF: Von Willebrand factor, PS: phosphatidylserine, ICAM-2: intracellular cell adhesion molecule, PSGL-1: P-selectin glycoprotein ligand-1. This image was created with BioRender (https://biorender.com/).

### Serum Amyloid A

SAA was also significantly upregulated in our Long COVID samples. SAA contains binding sites for the <u>extracellular matrix</u> components, <u>laminin</u> and heparin/heparan sulfate, as well as the RGD-like (arginine-glycine aspartic acid) adhesion motif (*58*). Immobilized SAA also binds to the αIIbβ3 receptor on platelets (*58*) and may also directly bind to toll-like receptors (TLR) on endothelial cells (*68*). Dysregulated SAA is associated with endothelial dysfunction and early-stage atherogenesis (*69*). SAA binding to on endothelial cells results these cells express ICAM-1, VCAM-1, E-selectin, TNF-α, IL-1, and IL-6, together with pro-thrombotic factors (*68*) (see Figure 7).

**Figure 7:**
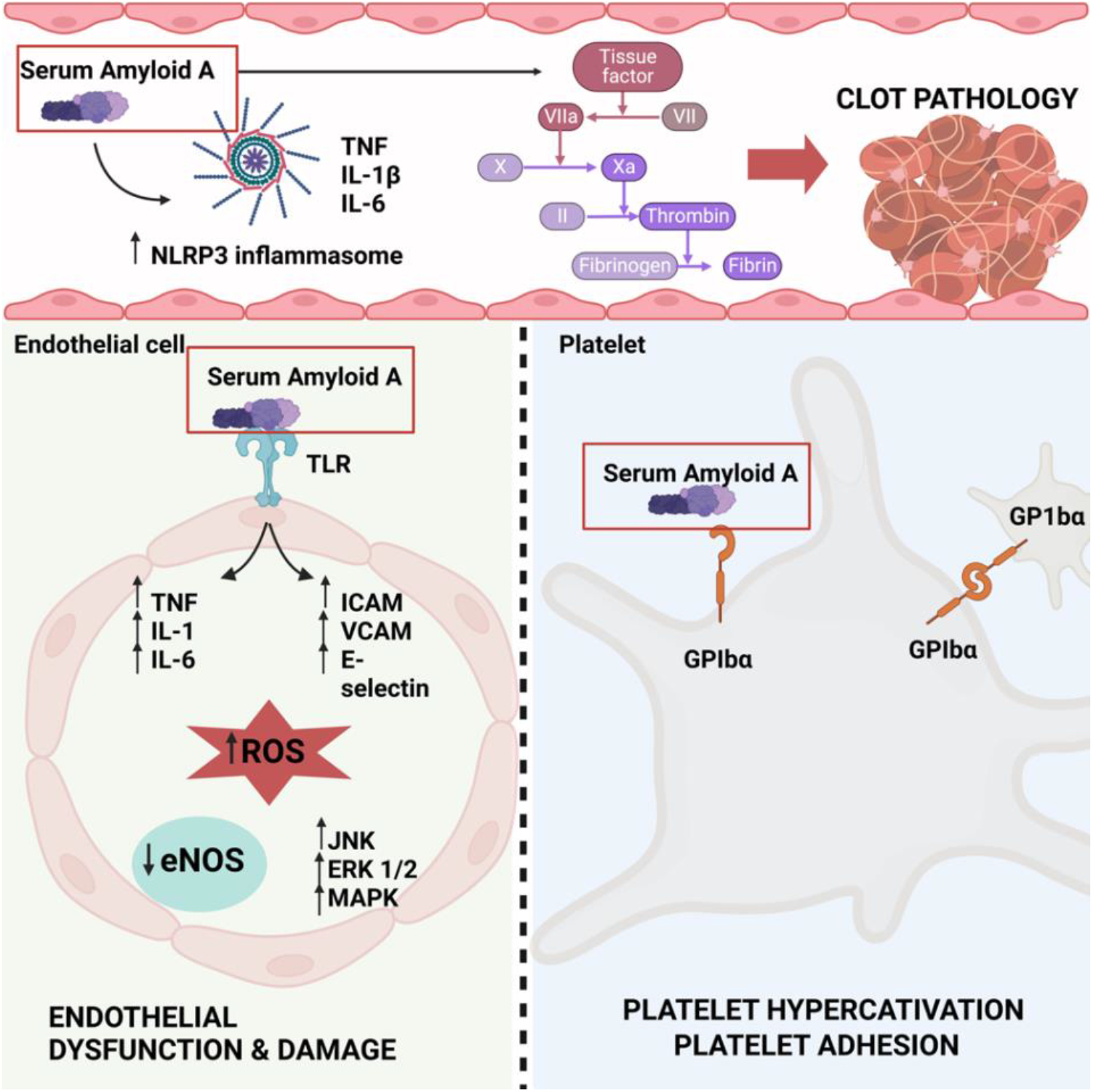
Serum Amyloid A and its receptor and signalling in endothelial cells and platelets. In circulation, SAA activates the NLRP3 inflammasome, promoting the production of inflammatory molecules. SAA also directly promotes TF, that activates the coagulation cascade to produce thrombin. SAA directly binds to endothelial cells via the Toll-like receptor. This instigates the increase of inflammatory molecules and adhesion molecules that ultimately increases the production of ROS. When ROS is increased in endothelial cells, it downregulates eNOS and upregulates JNK, ERK1/2, and MADK. This sequence of events may cause endothelial dysfunction and damage. Alternatively, SAA bind to the GP1bα receptor on platelets and cause platelet hyperactivation and platelet adhesion. Endothelial damage and platelet hyperactivation are also directly involved in coagulation pathology. **Abbreviations:** SAA: Serum Amyloid A, NLRP3: NLR family pyrin domain containing 3, TLR: toll like receptor, TNF: tissue necrosis factor, IL: interleukin, ICAM: Intercellular adhesion molecule-1, VCAM: Vascular cell adhesion molecule, ROS: reactive oxygen species, eNOS: endothelial nitic oxide synthase, JNK: Jun N-terminal kinase, ERK: extracellular receptor kinase, MAPK: Mitogen-activated protein kinases, GPIbα: platelet glycoprotein Ib alpha. This image was created with BioRender (https://biorender.com/).

### E-selectin

We also found that E-selectin is significantly upregulated in serum from individuals with Long COVID. This is a notable finding as E-selectin is typically undetected in endothelial cells that have not yet been activated (*70*). Upon inflammatory stimulation, E-selectin acts as an adhesion molecule and tethers leukocytes to endothelial cells (*71*) through supporting the initial rolling of leukocytes on activated endothelium (*72*). Several adhesion molecules including E-selectin, are induced and upregulated on endothelial cells during inflammation (*49*). The rolling of leukocytes on E-selectin engages P-selectin glycoprotein ligand-1 (PSGL-1) and cluster of differentiation 44 (CD44) to initiate signalling through a common pathway that requires lipid rafts, the cytoplasmic domain of PSGL-1, all 3 Src family kinases (SFKs), the immunoreceptor tyrosine-based activation motif (ITAM) adaptors, DNAX activating protein of 12 kDa (DAP12) and Fc receptor (FcR) y, the Tec family kinase Btk, and lastly, p38 (*75*). This signalling cascade subsequently activates integrin leukocyte function-associated antigen 1 (LFA-1) to a conformation that enables slow rolling to occur (*75*). Rolling and adhesion of leukocytes due to E-selectin signals, result in an increase of CD11b/CD18 (Mac-1), ultimately indirectly facilitating binding of leucocyte s to platelets (*76*) (see Figure 8).

**Figure 8:**
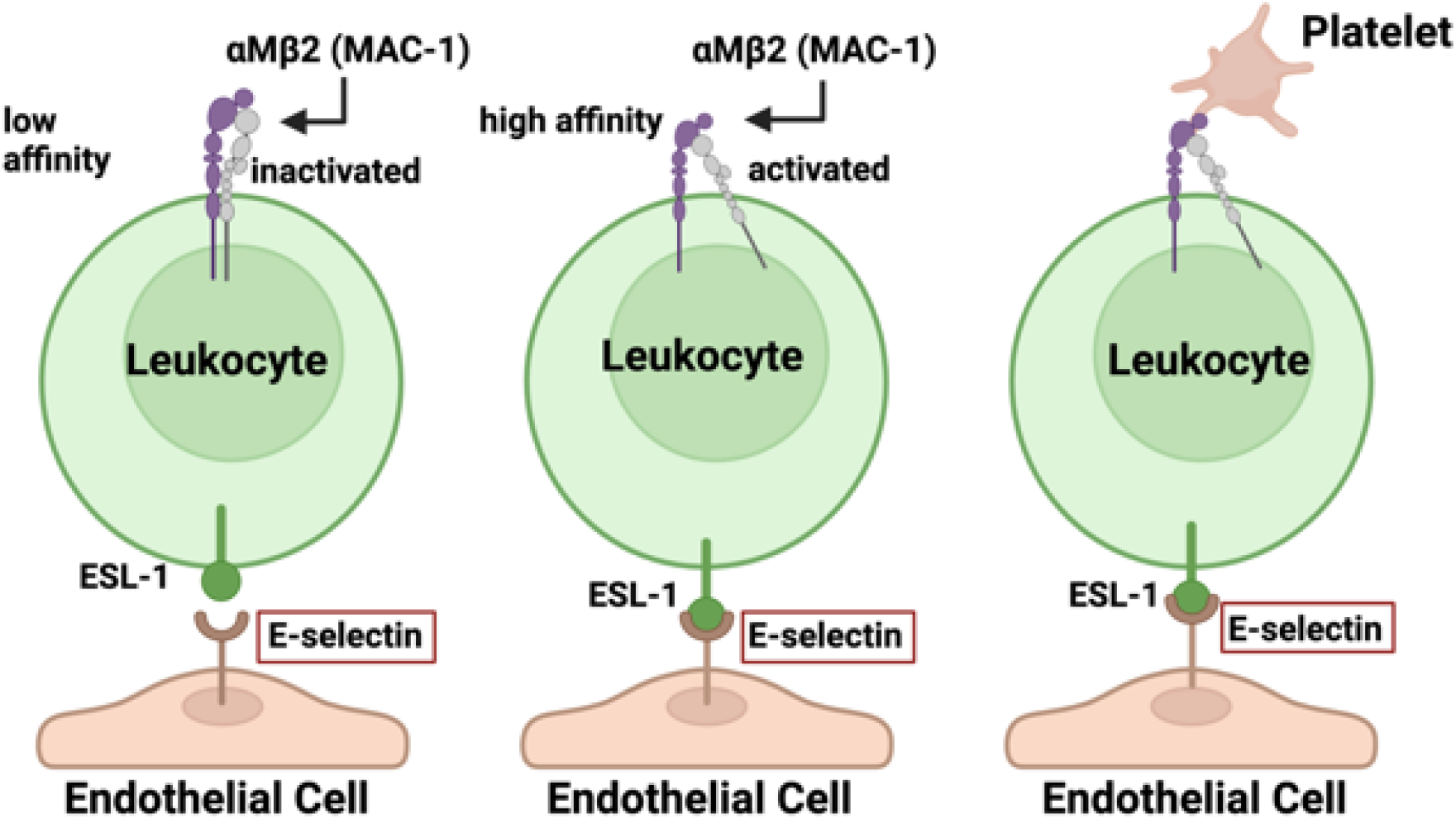
Mechanism by which E-selectin affects platelet adhesion to leukocytes (adapted from (*76*). Abbreviations: ESL-1, E-selectin ligand-1; MAC-1, macrophage-1 antigen. This image was created with BioRender (https://biorender.com/).

**Figure 9:**
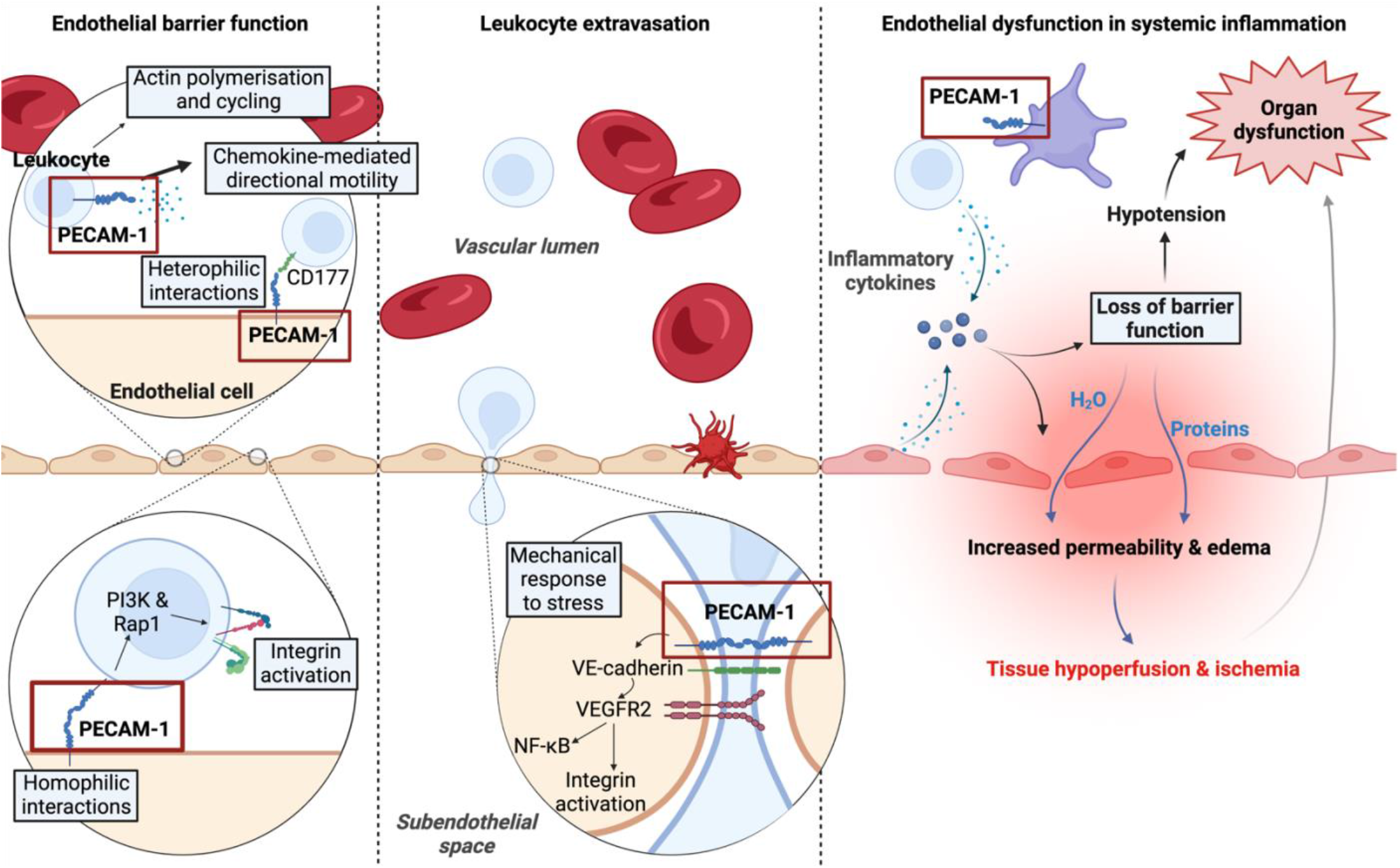
The functions of PECAM-1 in leukocyte binding and motility as well as the regulation of vascular permeability and maintenance of the endothelial barrier functioning (adapted from (Privratsky et al., 2010) and (Hellenthal et al., 2022) **Abbreviations:** PECAM-1, platelet endothelial cell adhesion molecule-1; Rap1, repressor/activator protein 1; PI3K, phosphatidylinositol-3-kinase; VE-cadherin, vascular endothelial-cadherin; VEGFR2, vascular endothelial growth factor receptor 2; NF-κB, nuclear factor-kappa B. This image was created with BioRender (https://biorender.com/).

### PECAM-1

We also found a significant upregulation of PECAM-1 in serum from individuals with Long COVID. PECAM-1 has been found to be expressed on endothelial cell membranes, platelets, and leukocytes (*51, 77-81*). In endothelial cells, PECAM-1 signals through the Syk/PECAM-1 signaling pathway (*79*) as a response to shear stress, causing tension changes across the junctional proteins VE-cadherin and PECAM-1. Onset of flow results in decreased force across VE-cadherin, and a simultaneous increase in force across PECAM-1 (*82*). The changes in tension across VE-cadherin and PECAM-1, activate vascular endothelial growth factor receptor 2 (VEGFR2) (*82*), followed by PI3K activation and the transcription of pro-inflammatory genes through the NF-κB pathway (*83*).

Even though PECAM-1 is known to be expressed on platelets (*81*), its role in platelets is still unclear. Unlike the other inflammatory markers, PECAM-1 is thought to function as a selective inhibitor for platelet aggregation and signalling (*84*). Mice models with decreased platelet-derived PECAM-1 showed increased responsiveness to collagen stimulation and led to larger thrombus formation in vitro, further implicating PECAM-1 in the inhibition of platelet function (*81*). In addition, PECAM-1 has shown to inhibit platelet protein tyrosine phosphorylation stimulated by thrombin, leading to inhibition of calcium mobilisation from intracellular stores (*81, 84*).

Apart from showing that both E-selectin and PECAM-1 are upregulated in blood from individuals with Long COVID, we were also able to induce significant microclot formation and platelet hyperactivation when adding these two purified molecules to healthy samples. These results suggest that when both E-selectin and PECAM-1 are in circulation, they may interact with soluble plasma proteins by engaging in direct protein-protein interactions. This protein-protein interaction results in misfolding of the plasma proteins and microclot formation. Both the molecules were also found to trigger platelet activation, suggesting direct platelet receptor-molecule interactions. These results further provide evidence of significant interactions between circulating inflammatory molecules in individuals with Long COVID.

### α-2-antiplasmin

We have previously found that α-2AP is significantly upregulated inside microclots found in samples from individuals suffering from Long COVID (*10, 19*). Here we also report a significant upregulation of α-2AP in the soluble part of the blood. This antifibrinolytic protein covalently binds to plasmin, and in so doing inhibits its function to solubilize fibrin polymers (*85*). When α-2AP is increased in circulation and comes in contact with plasmin, the C-terminal lysine residue of α-2AP binds non-covalently to the kringle domains of plasmin and forms a 1:1 stoichiometric complex (*86*). Increased levels of α-2AP therefore prevent the fibrinolytic system from dissolving pathologic thrombi resulting in venous thrombosis, pulmonary embolism, arterial thrombosis, and ischemic stroke (*43*) (see Figure 10).

**Figure 10:**
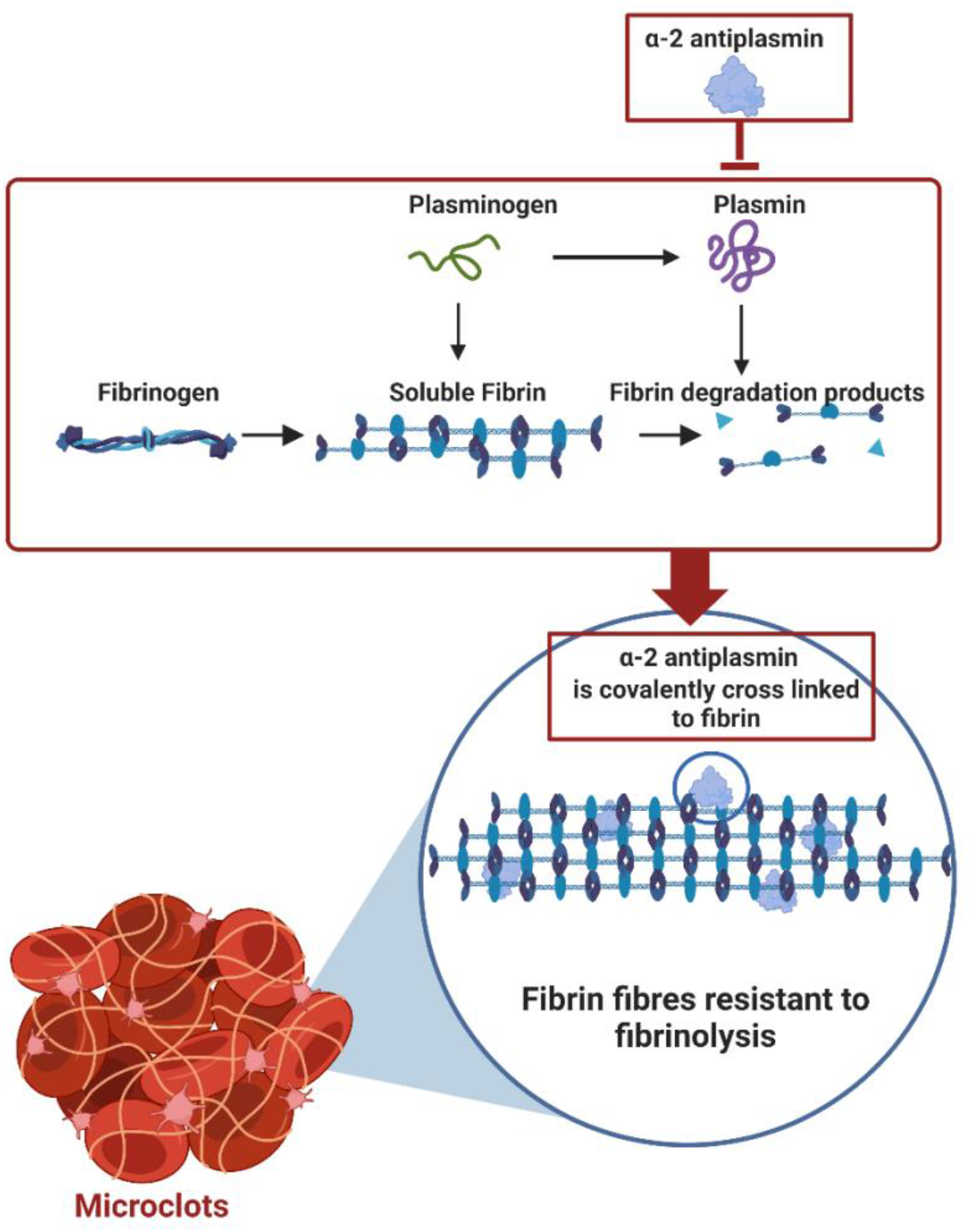
Alpha 2 antiplasmin binds covalently to plasmin, thereby inhibiting its fibrinolytic properties. Alpha 2 antiplasmin also covalently cross links to fibrin fibres within the thrombus. Collectively, this endorses insoluble fibrin fibres that ultimately causes insoluble microclots. This image was created with BioRender (https://biorender.com/).

## CONCLUSION

Dysregulation of coagulation, and in particular the formation of fibrin amyloid microclots that are resistant to proteolysis and can then cause ischaemia-reperfusion injury (*9, 11*), is a key part of the pathogenesis of Long COVID. In fact, we would argue that whilst there may be many mechanisms at play in the pathogenesis of Long COVID, the common pathological process is *thrombotic endotheliitis*. This may be characterised by the formation of anomalous fibrinaloid microclots, hyperactivated platelets, endotheliitis and elevated levels of prothrombotic inflammatory molecules which interact with each other as well as with platelets and the endothelium.

In this paper we have demonstrated significantly increased concentrations of VWF, PF4, SAA, α-2AP, E-selectin, and PECAM-1 in the soluble fraction of blood from individuals with Long COVID when compared to those without Long COVID. Each of the individual molecules can cause significant endothelial dysfunction and platelet activation, ultimately resulting in severe clotting imbalance and endothelial pathology. Figure 11 provides an overview diagram of the receptors and signalling pathways of our chosen molecules.

**Figure 11:**
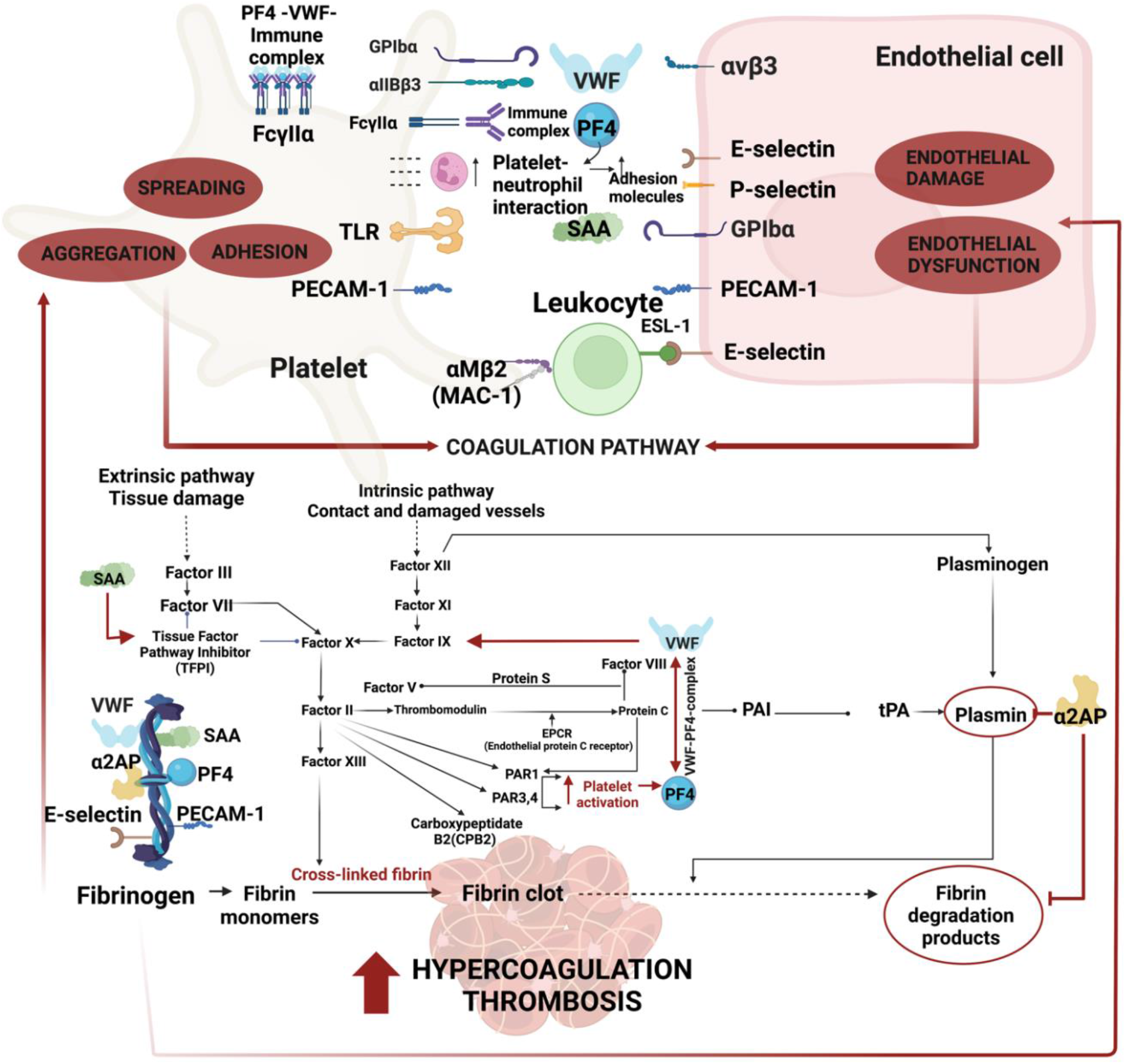
Overview diagram of the receptors and signalling pathways of Von Willebrand Factor (VWF), Platelet Factor 4 (PF4), Serum Amyloid A (SAA), α-2 antiplasmin (α-2AP), endothelial-leukocyte adhesion molecule 1(E-selectin) and Platelet endothelial cell adhesion molecule (PECAM).

Figure 12 provides an overview of the vascular damage these molecules may cause when in circulation.

**Figure 12:**
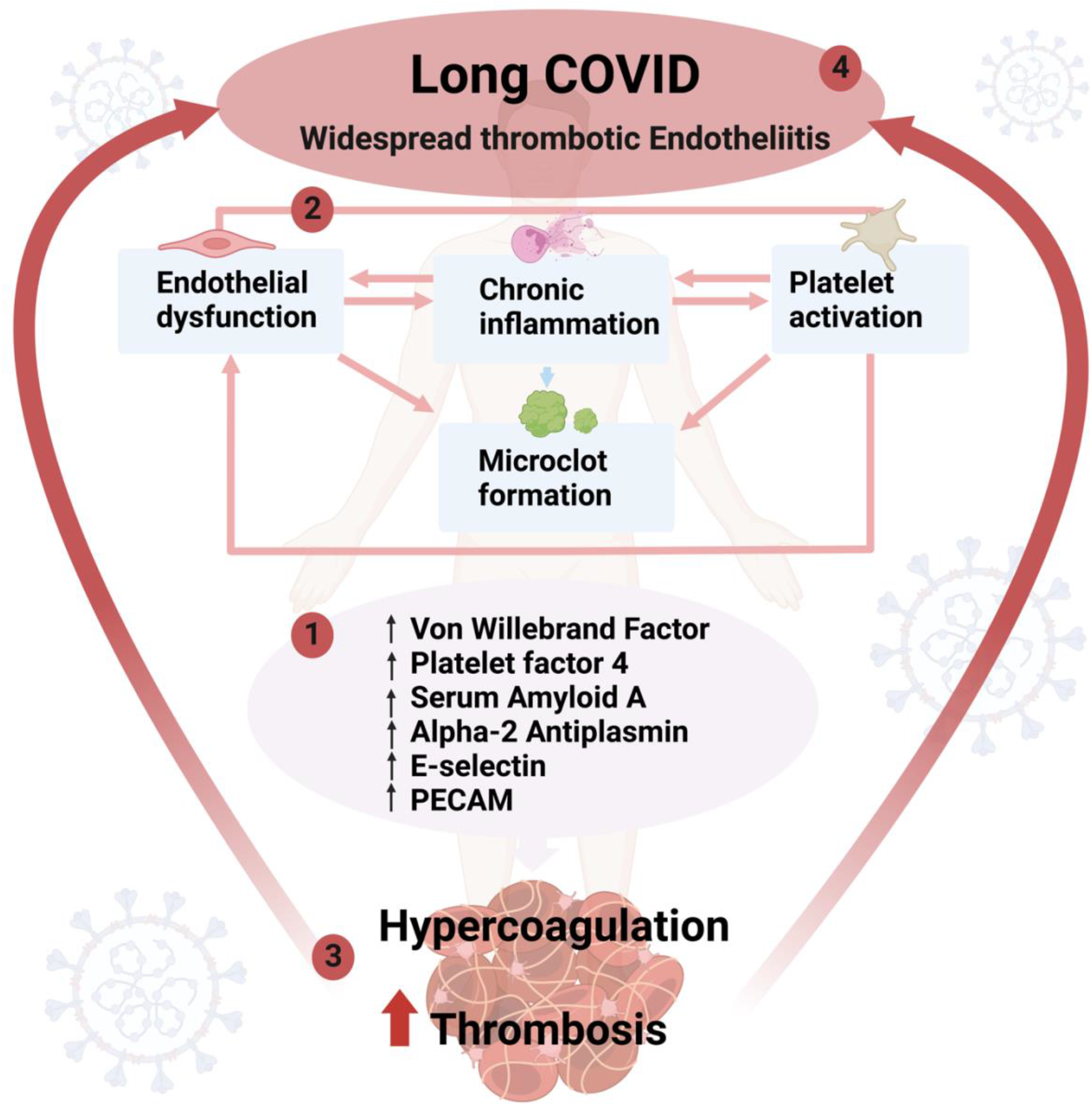
Overview of pathologies in Long COVID that develop as a direct response to the presence of **1)** the suit of six molecules (Von Willebrand Factor (VWF), platelet factor 4 (PF4), serum amyloid A (SAA), α-2-antiplasmin (α-2AP), PECAM-1 and E-selectin) that we propose as markers to identify **2)** endothelial damage, chronic inflammation and platelet activation, that drives **3)** clotting and ultimately is central in **4)** widespread thrombotic endotheliitis individuals with Long COVID.

Up to now, we have demonstrated significant microclot formation and platelet activation in all Long COVID patients whom we have studied using fluorescence microscopy (*2*), as well as detailed proteomics analysis revealing several-fold increased concentrations of inflammatory molecules entrapped within the microclots. Unfortunately, both fluorescence microscopy and proteomics are complex techniques usually only available in research facilities and are not currently available in clinical laboratories.

In the current paper, whilst only the mean level of alpha-2-antiplasmin exceeded the upper limit of the laboratory reference range in Long COVID patients, it was noteworthy that the average levels of the other 5 molecules measured were significantly higher in Long COVID patients as compared to the controls-even though technically they were within the ‘normal’ range. This is alarming if we take into consideration that a significant proportion of the total burden of these inflammatory molecules is entrapped inside fibrinolysis-resistant microclots (*2, 10, 17*).

Given the significant increase in the rates of thrombotic events following even a mild COVID-19 infection (*7*), it is imperative that the thrombotic endotheliitis is treated urgently. There is an immediate need for a microclot detection method that can be rolled out for clinical use. We argue that such a microclot detection method together with the demonstration of relatively high levels of the six molecules described in this paper, would be good evidence for an ongoing thrombotic endotheliitis. The optimal therapeutic regimen is yet to be defined; however given the complex pathophysiology, such therapy is likely to incorporate antiplatelet drugs, as well as agents acting on the enzymatic pathway of coagulation.

## Data Availability

All data produced in the present study are available upon reasonable request to the authors

## Author Contributions

“Conceptualization, E.P. and D.B.K.; Methodologies, S.T., E.P., C.N., T.U. Sample curation, C.V. Clinician input: G.J.L. A.K. and M.A.K.; E.P. and S.T. writing of paper. All authors reviewed and edited the paper.

## Institutional Review Board Statement

The study was conducted in accordance with the Declaration of Helsinki, and approved by the Institutional Review Board of Stellenbosch University (B21/03/001_ COVID-19, project ID: 21911 (Long COVID registry) and N19/03/043, project ID 9521 with yearly re-approval).

## Informed Consent Statement

Informed consent was obtained from all subjects involved in the study.

